# Psychometric properties of the Depression Stigma Scale in the Portuguese population and its association with gender and depressive symptomatology

**DOI:** 10.1101/2020.09.14.20194167

**Authors:** Virgínia da Conceição, Inês Rothes, Milton Severo, Kathleen Griffiths, Ulrich Hegerl, Ricardo Gusmão

## Abstract

**Background:** Stigma is one of the most significant constraints on people living with depression. There is a lack of validated scales in Portugal to measure depression stigma; therefore, validation of the Depression Stigma Scale (DSS) is an essential step to the depression stigma research in Portugal.

**Methods:** We developed the adaptation process with the ITC Guidelines for Translation and Adapting Tests taken into consideration. We collected the sample as part of the OSPI program – Optimizing suicide prevention programs and their implementation in Europe, specifically within the application in Portugal, and included 1693 participants. Floor-ceiling effects and response ranges were analyzed, and we calculated Cronbach alphas, conducted a Principal Component Analysis and Confirmatory Analysis. Validity evidence was tested with two well-documented hypotheses, using data on gender and depression symptoms.

**Results:** The sample was well comparable with the general Portuguese population, indicating its representativeness. We identified a three-factor structure in each subscale (personal and perceived stigma): weak-not-sick, discrimination, and dangerous/unpredictable. The Cronbach’s alphas were satisfactory, and validity was confirmed.

**Conclusions:** This study established the validity and demonstrated good psychometric properties of the DSS in the Portuguese population. The validation of the DSS can be beneficial in exploring stigma predictors and evaluating the effectiveness of stigma reduction interventions.

## Introduction

The stigma associated with mental illnesses was the subject of extensive study in recent years. Two relevant concepts of stigma are social and self-stigma. Harmful attitudes and discriminatory behavior towards people with mental illness characterize social stigma (1). The internalization of stigmatizing beliefs typifies self-stigma by the person living with mental illness (1) which often causes deep feelings of shame and guilt and may compromise the help-seeking and treatment process (2). Much of the recent research has focused on two types of social stigma: personal stigma and perceived stigma (Griffiths et al., 2008). Personal stigma refers to one’s own beliefs about depression, and perceived stigma refers to one’s beliefs about the attitudes of others.

The apparent adverse effects of mental illness stigma described in the literature, such as the reinforcement of some pathological symptoms such as lack of self-esteem and social isolation (3), constraints on professional integration and access to mental health care (2, 4) have driven the stigma research from the general concept of mental illness to the specific stigma attributed to a particular diagnosis.

Stigma is one of the most significant constraints on people living with depression with the community and stigmatized institutional responses similar to those experienced by people with psychosis or chronic mental illness (5). A study from 2013 showed that 79% of the participants had experienced discrimination associated with their depressed condition, and between 20% and 37% have compromised their actions because of anticipated discrimination (3). Among people with common mental disorders in Europe, there is 14.8% of both embarrassment and discrimination experiences, being more common among individuals with lower education and those married (6).

In 2015, the estimated prevalence of depression worldwide was 4.4% (7). In contrast, in Portugal, the prevalence in the same year was estimated at 5.7% and is responsible for 8.5% of the total Years Lived with Disability (7), raising the importance of depression stigma research.

Even though there is an increasing number of researches about depression stigma associated, it is essential to widen the range of sociocultural contexts in which the studies are developed (8) and the adoption of an instrument used in a variety of countries would allow overcoming difficulties related to the methodological discrepancies between existing studies (9).

Currently, in Portugal, there is a lack of validated scales to measure depression stigma. Although data about depression stigma in Portugal has been published under the studies resulting from the OSPI research program (10, 11), and its psychometric properties of the instrument used were explored at the time, the validation of the Depression Stigma Scale (12) was not yet published and is an essential step to the depression stigma research in Portugal.

The Depression Stigma Scale was developed by the Centre for Mental Health Research at the Australian National University to measure stigma associated with depression. It has two subscales that measure two different types of stigma: personal and perceived. The Personal Stigma Subscale measures stigma in the respondent’s attitudes towards depression by indicating how strongly they agree with nine statements about depression. The Perceived Stigma Subscale measures the respondent’s perception about the attitudes of others towards depression by asking them to indicate what they think most other people believe about the same nine statements. Measurement of responses to each item are on a five-point scale (ranging from zero ‘strongly disagree’ to four ‘strongly agree’). Higher scores indicate higher levels of depression stigma.

The Depression Stigma Scale has been defined as an excellent instrument to measure attitudes towards depression (13) and has been used in recent studies in a wide range of cultures (14-17). The detailed study of its psychometric properties is of enormous value to the understanding and researching depression stigma and the study of the effects of anti-stigma campaigns and the development of effective stigma-reduction interventions.

## Methods

We took the ITC Test Translation and Adaptation Guidelines (18) into consideration in the scale adaptation process and the second edition (19) into account in the validation process.

We obtained permission from the original developer of the scale that actively collaborated in the adaptation process, being one of the authors of this work. Since the adaptation occurred during the Optimizing suicide prevention programs and their implementation in Europe (OSPI) (10), a group of experts concerning the cultural differences of the stigma construct was involved in the process of judgments of the construct-item match and on the evaluation of its suitability for the Portuguese language, as well as the evaluation of potential cultural differences.

### Adaptation process

The translation of the Depression Stigma Scale (DSS) Portuguese version followed the double-translation and reconciliation procedure: two independent researchers, Portuguese natives, translated the questions from English to Portuguese, then the differences in translation were analyzed and conciliated by a third Portuguese native depression stigma expert. We then asked a group of native Portuguese to review the scale to ensure that the test instructions and item content had the intended meaning, with particular account for the stigma context in Portugal. This last group of researchers was both specialists in depression as well as in stigma.

Since the DSS is a five-point Likert scale, with answers ranging in order of agreement, and this is a widespread method of scale construction in Portugal, the item format was considered suitable.

### Participants

The sample collection for this study was part of the OSPI program – Optimizing suicide prevention programs and their implementation in Europe, specifically within the application in Portugal (6, 20), and the current study assumed a cross-sectional study design.

The adopted methodology for sample collection was discussed in-depth and approved by all the project consortium.

Since there are no listings of mobile phones in Portugal, we extracted a selection of participants from the cable telephone network listed numbers, using the random digit dialing method to numbers belonging to Almada and Amadora municipalities. Trained interviewers conducted telephone contacts, and fieldwork was performed at the end of the day and on weekends to achieve data representativeness regarding gender and age.

Almada and Amadora combined have above 350 000 inhabitants, making them two of the most populous counties in Portugal. Due to its high urban population density and the socio-economic diversity of its inhabitants, these municipalities were considered representative of the Portuguese population (10, 11).

The participation rate was 46%, considering the response rate has been decreasing in the past decades (21), and the mean response rate in 2012 for telephone surveys was 30.2%, the sample consent bias was considered minimal.

Of the 2009 participants in the OSPI research program (Hegerl et al., 2009, Coppens et al., 2013), we excluded 316 participants from the current study due to their exposure to the OSPI intervention aimed to promote literacy on depression and, consequently, help-seeking behavior. Collection of data occurred between 2009 and 2010.

### Instruments

We asked participants to answer a questionnaire containing sociodemographic queries concerning the respondent’s sex, age, professional occupation, and the complete Depression Stigma Scale, as well as the Mental Health Inventory 5 (MHI-5) (22, 23) in order to screen for depressive symptoms.

The MHI-5 is a brief version of the 38-item MHI developed in 1983 (24) The MHI-5 was developed for its use with the general population, and it includes items on psychological well-being. The five 6-point Likert items access psychological well-being (2 items) and the absence of psychological distress (3 items).

### Data Analysis

In addition to computing a continuous score for each DSS, we transformed the DSS score into percentages, following the original scale (Griffiths et al., 2004), with higher percentages indicating greater stigma levels. The same procedure was carried out for the MHI-5 scale, transforming the scores in a 0 to 100 range; higher scores meant better mental health than lower scores.

In order to study the psychometric properties of the DSS, we analyzed floor-ceiling effects and response ranges, calculated Cronbach alphas, conducted a principal component analysis, and tested the Model Fit using Confirmatory Analysis.

Descriptive statistics of the DSS-Personal items were estimated, along with the corrected item-test correlation. We assessed the univariate normal distribution of the items using the Kolmogorov-Smirnov test and calculated Loadings with Promax with Kaiser normalization, considering Eigenvalue above 1.

To obtain principal component analysis, we used the Determinant, Kaiser-Meyer-Olkin, and Bartlett test, in the full scale, and both subscales: personal depression stigma and perceived depression stigma.

We examined the model fit using the Normed Fit Index (NFI), Comparative Fit Index (CFI), Adjusted Goodness of Fit (AGFI), Tucker-Lewis Index (TLI), Root Mean Square Residual (RMR), and Root Mean Square Error of Approximation (RMSEA). The criteria for an acceptable or good model fit were CFI, TLI, and NFI > 0.95, AGFI > 0.90, RMR < 0.06, and RMSEA < 0.08 were considered an acceptable model fit (Hu and Bentler, 1999).

We based validity evidence on the test of two of the better-documented hypotheses (25-27):

(1) women present lower scores of personal stigma and higher perceived stigma than men and

(2) depressive symptomatology is associated with higher personal and perceived stigma. We calculated means and standard deviations for continuous variables, student t-tests to examine the differences between sexes, and separate linear regression analyses to assess the effects of symptomatology and other factors on each depression stigma subscale, as well as Pearson correlations.

IBM SPSS Statistics, Version 24.0 software package was used to conduct statistical analysis, and IBM AMOS was used to conduct the Confirmatory Analysis.

### Ethical Considerations

Trained interviewers explained in the first contact the objectives of the study and obtained verbal informed consent. Anonymity and confidentiality of the data collected were guaranteed.

The approval of the study protocol occurred in the scope of the OSPI program (Coppens et al., 2013; Kohls et al., 2017). In Portugal, the OSPI study was approved by the Medical Sciences Faculty of the New University of Lisbon’s ethics commission in May 2009.

## Results

### Sample description

As to gender and age, 53.6% were women, and the mean age was 47.2 years (SD=18.17, range 18-90). National data estimates a mean age in the Portuguese population of 45.2, with a gender distribution of 52.2% women and 47.8% men. We found 8.6% of unemployment in our sample, whereas in 2009, the unemployment rate in Portugal was 9.4%.

Most of the individuals on our sample were married (52.9%), followed by single individuals (33.2%), widowed (7.1%), and divorced (6.8%). The marital status of the Portuguese population follows a similar distribution: 54.8% are married, 30.1% single, 8.5% widow, and 6.6% divorced.

### Depression Stigma Scale psychometric characteristics

As we can observe in Table 1, the KMO (Kaiser-Meyer-Olkin) test value is sufficiently close to 1, which indicates that the correlation patterns are relatively compact, so factorial analysis should produce factors other than reliability. The Bartlett test of sphericity shows that the correlation matrix is significantly different from the identity matrix, confirming that it is appropriate to perform the factor analysis. There were no missing values, and we used all response options in all items. Besides, there were no evident inconsistencies in the frequency of responses, and no ceiling or floor effects were detected.

**Table 1:** Depression Stigma Scale Portuguese version Psychometrics’ properties

| Analysis | Indicators | Results |  |
| --- | --- | --- | --- |
| Principal component analysis assumption verification | Determinant, Kaiser-Meyer-Olkin, and Bartlett test. | Total Scale | Determinant: 0.22; KMO>0.75; Bartlett test $p<0.01$ |
| | | Personal Subscale | Determinant: 0.27; KMO>0.70; Bartlett test $p<0.01$ |
| | | Perceived Subscale | Determinant: 0.18; KMO>0.80; Bartlett test $p<0.01$ |
| Sensitivity | Response frequencies, skewness, and kurtosis | Use of all response options and normal distribution of answers. No observation of a floor-ceiling effect. |  |
| Factorial Validity | Loadings with Promax with Kaiser normalization. (Eigenvalue above 1) | Two dimensions: Personal and perceived subscales. |  |
| Reliability | Cronbach's Alpha | Total scale: 0.75<br>Personal Subscale: 0.71<br>Perceived Subscale: 0.76 |  |
| Model Fit Analysis | NFI, AGFI, CFI and RMSEA | NFI= 0.914; AGFI=0.967; CFI=0.931; TLI: 0.901; RMR: 0.040; RMSEA=0.046 |  |

As we expected the items to be correlated, we used the Promax rotation with the Kaiser normalization. The criteria for the extraction was 50% of the total variance explained and an Eigenvalue higher than 1. We extracted two dimensions from the scale with these criteria: the first nine items corresponding to the personal subscale and the last nine items corresponding to the perceived subscale.

When analyzing the subscales separately, both the eigenvalues (eigenvalues >1) and the scree plot suggested a three-factor solution of the personal depression stigma subscale and the remaining five items loaded >0.40, with no cross-loadings >0.20. The three factors accounted for 55.29% of the variance: the first factor explained 29.64%, 13.21% by the second, and 12.44% by the third factor. We named the first and last factors after Zhu and colleagues’ designation (28): *weak-not-seek* in the first factor (items 1, 2, 3) and *discrimination* in the third factor (items 7, 8, 9). The second factor was labeled *Dangerous/ unpredictable*, following Boerema and colleagues designation (26), corresponding to items 4, 5, 6. Their correlation was 0.36 (p<0.001).

We identified the same factor solution in the perceived depression stigma subscale, with the three-factor solution accounting for 59.55% of the variance. In this case, the first factor (*weak-not-seek*, items 10, 11, 12) explained 11.31% of the variance, the second factor (*Dangerous/ unpredictable*, items 13, 14, 15) explained 14.28% of the variance, and the third factor (*discrimination*, items 16, 17, 18) explained 33.96%. Their correlation was 0.38 (p<0.001).

We did not observe floor-ceiling effects. The percentage of participants endorsing each item ranged from 6-81% and 10-94% for the Personal and Perceived subscales, respectively.

Both Subscales show good Cronbach’s Alphas that did not increase after removing any of the items.

As expressed in table 1, the Model Fit Analysis showed good results in all indicators.

### Validity evidence

In the personal stigma subscale, women presented a statistically lower mean score (M=38.29, SD=13.98) than men (M=41.02, SD=13.95), t_(1691)_=4.00, p<0.001. On the other hand, women obtained higher mean scores on the perceived subscale (M=56.53, SD=14.83) than men (M=55.38, SD=13.89). However, the difference was not significant (t_(1691)_=-1.64, p=0.10).

MHI-5 scores showed a negative correlation with the personal depression stigma (r=-0.07, p <0.01); still, the correlation with the perceived depression stigma subscale was not significant (r=0.02, p=0.30).

As we can see in table 2, we detected significant effects on personal stigma from gender, age, and depressive symptomatology. Both age and being a man have positive effects on personal stigma, increasing their score. On the other hand, better MHI-5 scores, translating into better mental health, had a negative effect on personal depression stigma, decreasing the score.

**Table 2:** Effects of gender, age, and MHI-5 on personal depression stigma scores

|  |  | **B** | **95% CI** | **t** | **P** |
| --- | --- | --- | --- | --- | --- |
| Woman | Ref. |  |  |  |  |
| Men |  | **3.37** | **2.30, 4.65** | **26.87** | **<0.001** |
| Age |  | **0.23** | **0.18, 0.27** | **165.52** | **<0.001** |
| MHI-5 |  | **-0.10** | **-0.17, -0.02** | **6.41** | **<0.05** |

β=beta regression coefficients, Ref.=Reference category Significant results are shown in bold.

On the perceived depression stigma, gender and age presented the opposite effect compared to the personal stigma: age and being a man decreased the perceived depression stigma score, as shown in table 3. MHI-5 scores did not produce a significant effect on perceived stigma scores.

**Table 3:** Effects of gender, age, and MHI-5 on perceived depression stigma scores

|  |  | **B** | **95% CI** | **t** | **P** |
| --- | --- | --- | --- | --- | --- |
| Woman | Ref. |  |  |  |  |
| Men |  | **-1.46** | **-2.82, -0.09** | **4.38** | **<0.05** |
| Age |  | **-0.01** | **-0.26, -0.08** | **30.04** | **<0.001** |
| MHI-5 |  | 0.03 | -0.05, 0.11 | 0.57 | 0.45 |

β=beta regression coefficients, Ref.=Reference category Significant results are shown in bold.

## Discussion

The present study is the first to examine the Depression Stigma Scale (DSS) psychometric properties in the Portuguese population, which can be a significant step towards the depression stigma research in Portugal.

Even though our sample presented a mean age five years older than the mean age of the Portuguese population, we can consider our sample representative due to the similarities in other sociodemographic characteristics, such as gender distribution, occupation, and marital status.

Overall, the Portuguese version of the DSS showed good psychometric properties, suggesting that it was an appropriate instrument for future studies in the Portuguese population. Although, internal consistency in the perceived subscale was lower than that obtained in its original form (α=0.88) (Griffiths et al., 2004), and in the Dutch version (α=0.82) (Boerema et al., 2016), it was nevertheless satisfactory. Internal consistency for the personal subscale was similar to that reported in previous studies.

We confirmed each subscale as a particular dimension from the full scale, and all the items in each subscale presented good loadings. Additionally, and in similarity with the structure identified by Boerema and colleagues (26), the scree plot of each subscale indicated a three-factor solution: *weak-not-sick* (items 1, 2, and 3 from the personal subscale and 10, 11, and 12 in the perceived subscale), *dangerous/unpredictable* (items 4, 5, 6 and 13, 14 and 15 form the personal and perceived subscale respectively) and lastly, *discrimination* (items 7, 8 and 9 in the personal subscale and 16, 17 and 18 in the perceived subscale). This structure showed good fit indices in the confirmatory factor analysis, an exciting finding since the scale structure seems to be influenced by cultural factors, ranging from a one-factor structure (29) to a three-factor (26).

In order to access validity evidence, we tested two well-documented hypotheses. The first one, widely observed in previous literature, stated that we expected to see women with lower personal stigma and higher perceived stigma than men. When analyzing mean differences between genders, the difference between scores in the personal scale was clear: women showed lower personal stigma than men in agreement with the previous literature (12, 25-27); however, we observed no difference in the perceived depression stigma subscale. Differences in the perceived stigma have not been as consensual in the literature as those observed in the personal stigma. On the one hand, we can find research that supports that women present higher perceived stigma than men (27), in the other, we find literature supporting the absence of differences (30). Even though in the direct comparisons we found no statistically significant difference between genders in the perceived scale, in the regression, gender has shown to have a significant effect on perceived depression stigma scores, which can indicate that there are, in fact, gender differences, however subtle, and conditioned by other variables.

The second hypothesis led us to expect greater personal and perceived stigma in the population with the higher depressive symptoms. While, in our sample, we confirmed the hypotheses for personal stigma, in the perceived depression stigma, depressive symptoms did not show significant effects. We can hypothesize that perceived depression stigma can be more sensitive to other variables such as the proximity of the people participants are thinking when evaluating stigma around them (31) and previous experiences of help-seeking and close ones living with depression (1, 32).

One limitation of this study is related to the data collection because interviews were carried out by phone, probably delivering some desirable social answers. Another limitation is the absence of convergent validity of the scale, and future research is needed to examine the convergent validity in the Portuguese population.

Nevertheless, the scale presented good psychometric properties in the Portuguese population, and its validity was confirmed. Considering the well-recognized adverse effects of stigma, development of many initiatives aimed to reduce depression stigma in the populations, the existence of a validated scale can be crucial in evaluating the effectiveness of the interventions. Also, access to a validated scale allows us to explore better the depression stigma predictors and their effects on help-seeking and mental health promotion behaviors.

## Data Availability

Data avalabe upon request to the corresponding author

